# Asthma and COVID-19 in children – a systematic review and call for data

**DOI:** 10.1101/2020.05.04.20090845

**Authors:** Jose A. Castro-Rodriguez, Erick Forno

**Affiliations:** Division of Pediatrics, School of Medicine, Pontificia Universidad Católica de Chile, Santiago, Chile; Division of Pulmonary Medicine, Department of Pediatrics, University of Pittsburgh School of Medicine, Pittsburgh, Pennsylvania, US

## Abstract

**Rationale:** Whether asthma constitutes a risk factor for COVID-19 is unclear.

**Methods:** We performed a systematic literature search in three stages: First, we reviewed PubMed, EMBASE and CINAHL for systematic reviews of SARS-CoC-2 and COVID-19 in pediatric populations, and reviewed their primary articles; next, we searched PubMed for studies on COVID-19 or SARS-CoV-2 and asthma/wheeze, and evaluated whether the resulting studies included pediatric populations; lastly, we repeated the second search in BioRxiv.org and MedRxiv.org to find pre-prints that may have information on pediatric asthma.

**Results:** In the first search, eight systematic reviews were found, of which five were done in pediatric population; after reviewing 67 primary studies we found no data on pediatric asthma as a comorbidity for COVID-19. In the second search, we found 25 results in PubMed, of which five reported asthma in adults, but none included data on children. In the third search, 14 pre-prints in MedRxiv were identified with data on asthma, but again none with pediatric data. We found only one report by the U.S. CDC stating that 40/345 (~11.5%) children with data on chronic conditions had “chronic lung diseases including asthma”.

**Conclusion:** There is scarcely any data on whether childhood asthma (or other pediatric respiratory diseases) constitute risk factors for SARS-CoV-2 infection or COVID-19 severity. Studies are needed that go beyond counting the number of cases in the pediatric age range.

## INTRODUCTION

The current outbreak of coronavirus disease 2019 (COVID-19), caused by the severe acute respiratory syndrome coronavirus-2 (SARS-CoV-2), started in or around December, 2019, in Wuhan[1]. On January 30^th^, 2020 the World Health Organization (WHO) declared COVID-19 a pandemic health emergency[2]. Since then, COVID-19 has continued to spread quickly and has now become the most dangerous pandemic in over 100 years.

An interactive real-time COVID-19 reporting system set up by the Center for Systemic Science and Engineering at Johns Hopkins University[3] shows, as of the time of this writing, more than 3.1 million confirmed cases and over 217,000 deaths worldwide (led by the U.S., with one third of all cases and one quarter of all deaths). Globally, this corresponds to a ~7% case fatality rate, although rates vary widely among countries and subpopulations.

The first pediatric case in the literature was reported on January 2020 in 10-year-old boy from Shenzhen, China, whose family had visited Wuhan[4]. All epidemiological evidence to date suggests that SARS-CoV-2 infection is less severe in children than in adults. In the latest and largest study in the UK, including 16,749 patients hospitalized for COVID-19, only 239 (2%) were <18 years of age (including 139 who were <5 years old)[5]. Large studies in Italy and China have also shown very low case-fatality rates in children and adolescents[6]. Understandably, most studies have focused on adult populations, with very few studies and reviews in children. Moreover, accumulating data points to risk factors for severity and mortality in adults (e.g. older age, cardiovascular disease, diabetes, cancer, immunosuppression, obesity, tobacco smoking, etc)[5, 6], but there is very scarce evidence on whether or which risk factors exist in children. While COVID-19 is a multi-system disease, it predominantly affects the lungs, and thus it is critically important to understand whether chronic lung diseases place children at higher risk.

The main objective of this study was to identify whether asthma, the most common chronic respiratory disease in children, is a risk factor for SARS-CoV-2 infection or COVID-19 severity in the pediatric population.

## METHODS

We performed a systematic literature research in three stages: First, we searched PubMed, EMBASE and CINAHL using the terms “SARS-CoV-2 OR COVID-19” AND “systematic review” AND “children 0-18 years of age” to find systemic reviews on the topic, and then reviewed the primary studies included in those reviews. Second, we searched PubMed for “COVID-19 OR SARS-CoV-2” AND “asthma OR wheezing”, to directly find any studies on asthma/wheezing and COVID-19, and evaluated whether they included pediatric populations. Third, we repeated search #2 in BioRxiv.org and MedRxiv.org to evaluate whether existing pre-prints may have relevant pediatric asthma information. The last update of the searches was on May 1, 2020.

Both authors (JCR, EF) independently screened and retrieved articles. The same investigators independently assessed full texts of those primary studies included in the systematic review identified. Any discrepancies were resolved by discussion and consensus. If sufficient studies with relevant data were found, the plan was to perform a meta-analysis by asthma status.

## RESULTS

After removing duplicates, the first search yielded eight systematic reviews[5, 7-13]. Three of them were eliminated because they did include information on clinical characteristics in children[7-9]. Therefore, we evaluated five systematic reviews done at different periods during the pandemic and thus including somewhat different primary studies[10-13]. Castagnoli et al.[10] included 18 articles, 17 from China and 1 from Singapore (444 patients <10 years old and 553 aged 10-19 years), published up to March 3, 2020. Choi et al.[11] included 7 articles from China (225 pediatric patients) up to March 12, 2020. Chang et al.[12] included 9 studies from China (93 pediatric patients) up to March 15, 2020. The review by Ludvigsson[13] included 45 studies from China (the total number of patients was not described) up to March 19, 2020. And Streng et al.[14] included 8 studies from China (ranging from 6 to 2,143 patients) and one survey from Germany (33 patients) in hospitalized children, up to March 31, 2020. After excluding duplicates, we identified and reviewed 67 primary studies included in those five reviews.

None of the primary studies reviewed reported asthma or recurrent wheezing as a comorbidity or risk factor for COVID-19. Instead, some of those studies reported young age (especially children <1 year of age) as a group with more severe COVID-19. One large Chinese study[15] reported non-respiratory chronic conditions (hydronephrosis, leukemia receiving chemotherapy, and intussusception) among the 3 children required ICU support and mechanical ventilation all had coexisting conditions; one death occurred in the 10-month child with intussusception. Another study reported a patient develop shock with metabolic acidosis requiring ICU[16]; while a report from China[17] described one patient aged 10-19 years who died, without other clinical information, that probably is the same death in a 14-year-old boy described by Dong and colleagues[18]. Unfortunately, the two larger studies in Chinese pediatric patients, Dong et al.[18] (2,413 children) and Wu and MacGoogan[19] (965 children) did not report enough clinical data to identify comorbidities or risk factors for COVID-19 severity. In the German survey of 33 hospitalized children, 4 out of 22 (18%) children with clinical information had “respiratory comorbidities” without further details[14].

Our second search yielded 25 results in PubMed. Of those, four were primary studies that reported on asthma in adults[20-23]; one other was a guidance statement[24] that referenced a primary report that also included information on asthma in adults[25]. No studies from that search included information on asthma in children, although one case series reported two young children (ages 2 and 3 years) with history of atopic dermatitis and allergic rhinitis, who were hospitalized with COVID-19; both patients recovered[26]. Our third search yielded 26 pre-prints in BioRxiv and 86 in MedRxiv. None of the BioRxiv posts were relevant to our topic. Of the 86 pre-prints in MedRxiv, 14 non-duplicate studies included information on asthma[27-40], but none of them included specific information in children.

More recently, the CDC published a Morbidity Mortality Weekly Report (MMWR)[41] that included information from 2,572 U.S. children aged < 18 years. Of those cases, 345 had data on clinical and underlying conditions, and 80 of those children (23%) had at least one underlying condition. The most common underlying conditions were “chronic lung diseases (including asthma)” in 40 children, cardiovascular disease in 25, and immunosuppression in 10; separate information on asthma was not provided. Among the 295 cases for which data on both hospitalization status and underlying medical conditions was available, 28/37 (77%) hospitalized patients had one or more underlying medical condition (including all six patients admitted to an ICU); compared to 30/258 (12%) patients who were not hospitalized[41].

## DISCUSSION

In a systematic review of the literature, we only found one report described asthma/recurrent wheezing as a potential risk factor for COVID-19 in children. Importantly, none of the largest epidemiological studies including children with COVID-19 reported clinical findings or underlying characteristics to help assess whether asthma –or other chronic lung diseases–constitutes a risk factor for SARS-CoV-2 infection or COVID-19 severity.

COVID-19 affects primarily the lungs, and accordingly several international guidelines have designated some respiratory conditions as a potential risk factor for severe disease. Chinese guidelines[42] state that “children with a history of contact with severe 2019-nCoV infected cases, or with underlying conditions (such as congenital heart disease, bronchial pulmonary hypoplasia, respiratory tract anomaly, with abnormal hemoglobin level, severe malnutrition), or with immune deficiency or immunocompromised status… may become severe cases”. A recent statement from the EAACI Section on Pediatrics[24] declared that “patients with asthma (particularly severe or uncontrolled asthma) and immunodeficiency have also been classified to be at increased risk of developing severe COVID-19, based more on common sense rather than mounting evidence”. The Global Initiative for Asthma (GINA) recommends avoiding the use of nebulizers due to the increased risk of disseminating COVID-19 to other patients and healthcare staff; they thus recommend the use of pressurized metered dose inhalers (pMDI) as the preferred delivery system during asthma attacks[43]. A recent randomized controlled trial (RCT)[44] showed that even in children with severe asthma exacerbations, administration of albuterol/salbutamol and ipratropium by MDI with valved-holding chamber and mask along with oxygen by nasal cannula was more effective than nebulized administration. GINA[43] and the British Thoracic Society[45] do not recommend stopping oral steroids in the patients already taking them for asthma management, and they do not recommend avoiding them for acute asthma attacks even if due to COVID-19. The U.S. CDC, the Canadian Pediatric Society, and other professional associations have issued guidance for patients with asthma and/or allergies[46-48]. Other professional organizations, such as the American Academy of Pediatrics and the American Thoracic Society, have published interim guidelines that do not specifically address asthma, likely given a paucity of evidence[49, 50].

Rather than a risk factor, a recent review of data in adults found it that both asthma and COPD appear to be under-represented in the comorbidities reported for patients with COVID-19, compared with global estimates of prevalence for these conditions in the general population. This was in contrast to the prevalence of other chronic diseases such as diabetes, which occurred with higher frequency among patients with COVID-19 than the estimated national prevalence[51]; but consistent with several studies that have shown lower-than-expected prevalence of asthma among cases of COVID-19[20-23, 25]. If true, this could be due to several factors, including changes in the immune response or decreased risk secondary to chronic medications such as inhaled corticosteroids (ICS). In-vitro models have shown that ICS may suppress both coronavirus replication and cytokine production[52, 53]. And a recent study (in children and adults) showed that patients with asthma and respiratory allergies had reduced angiotensin-converting enzyme 2 *(ACE2)* gene expression in airway cells, suggesting a potential mechanism of reduced COVID-19 risk[54]. This is particularly noteworthy considering that one of the potential explanations for children being generally less affected than adults is the hypothesis that children have lower ACE2 receptor expression in alveolar type 2 cells[55]. However, the lower prevalence of asthma among COVID-19 cases could also stem from bias due to underdiagnosis and under-reporting, or because patients with chronic lung diseases may be especially cautious in practicing physical distancing and other measures to avoid infection. Finally, it is also conceivable that some milder cases of COVID-19 might be confused with exacerbations of respiratory disease, and/or that these patients may be reluctant to seek medical care even when sick and are thus never counted.

## CONCLUSIONS

After an extensive review of the current literature, only one study reported information on asthma as a potential risk factor for COVID-19 severity –but not mortality– in children. However, the largest studies to date have been limited to a description of the number of cases by age group, and so it remains unclear whether childhood asthma –or other pediatric respiratory diseases– are associated with COVID-19 risk or severity.

We hereby ask the public health community to move beyond confirming what’s already known –that the disease affects children and young adults less frequently and severely than older groups– and to study affected pediatric populations in more detail. Does asthma constitute a risk factor for COVID-19 in children? Do asthma severity or control modify the course of the disease? Are asthma medications (particularly ICS and systemic steroids) protective or detrimental? Given the limited numbers of pediatric cases in any one given center/country, collaborative international efforts may be the only way to shed light on the topic. This will be true not just for childhood asthma but for pediatric diseases in general.

## Data Availability

This manuscript is a review of publicly available literature.

## Notes

### Competing Interest Statement

The authors have declared no competing interest.

### Funding Statement

Dr. Forno’s contribution was partly funded by grant HL149693 from the U.S. National Institutes of Health (NIH).

## REFERENCES

1. Guan, W.J., et al., Clinical Characteristics of Coronavirus Disease 2019 in China. N Engl J Med, 2020. 382(18): p. 1708–1720.

2. Organization, W.H. WHO Director-General’s opening remarks at the media briefing on COVID-19: 11 March 2020. 2020 05/01/2020]; Available from: https://www.who.int/dg/speeches/detail/who-director-general-s-opening-remarks-at-the-media-briefing-on-covid-19---11-march-2020.

3. Dong, E., H. Du, and L. Gardner, An interactive web-based dashboard to track COVID-19 in real time. Lancet Infect Dis, 2020.

4. Chan, J.F., et al., A familial cluster of pneumonia associated with the 2019 novel coronavirus indicating person-to-person transmission: a study of a family cluster. Lancet, 2020. 395(10223): p. 514–523.

5. Docherty, A.B., et al., Features of 16,749 hospitalised UK patients with COVID-19 using the ISARIC WHO Clinical Characterisation Protocol. medRxiv, 2020: p. 2020.04.23.20076042.

6. Onder, G., G. Rezza, and S. Brusaferro, Case-Fatality Rate and Characteristics of Patients Dying in Relation to COVID-19 in Italy. JAMA, 2020.

7. Di Mascio, D., et al., Outcome of Coronavirus spectrum infections (SARS, MERS, COVID 1-19) during pregnancy: a systematic review and meta-analysis. Am J Obstet Gynecol MFM, 2020: p. 100107.

8. Viner, R.M., et al., School closure and management practices during coronavirus outbreaks including COVID-19: a rapid systematic review. Lancet Child Adolesc Health, 2020. 4(5): p. 397–404.

9. Chauhan, R.P., et al., Systematic Review of Important Viral Diseases in Africa in Light of the ‘One Health’ Concept. Pathogens, 2020. 9(4).

10. Castagnoli, R., et al., Severe Acute Respiratory Syndrome Coronavirus 2 (SARS-CoV-2) Infection in Children and Adolescents: A Systematic Review. JAMA Pediatr, 2020.

11. Choi, S.H., et al., Epidemiology and clinical features of coronavirus disease 2019 in children. Clin Exp Pediatr, 2020. 63(4): p. 125–132.

12. Chang, T.H., J.L. Wu, and L.Y. Chang, Clinical characteristics and diagnostic challenges of pediatric COVID-19: A systematic review and meta-analysis. J Formos Med Assoc, 2020.

13. Ludvigsson, J.F., Systematic review of COVID-19 in children shows milder cases and a better prognosis than adults. Acta Paediatr, 2020.

14. Streng, A., et al., [COVID-19 in hospitalized children and adolescents]. Monatsschr Kinderheilkd, 2020: p. 1–12.

15. Lu, X., et al., SARS-CoV-2 Infection in Children. N Engl J Med, 2020. 382(17): p. 1663–1665.

16. Chen, F., et al., [First case of severe childhood novel coronavirus pneumonia in China]. Zhonghua Er Ke Za Zhi, 2020. 58(3): p. 179–182.

17. Novel Coronavirus Pneumonia Emergency Response Epidemiology, T., [The epidemiological characteristics of an outbreak of 2019 novel coronavirus diseases (COVID-19) in China]. Zhonghua Liu Xing Bing Xue Za Zhi, 2020. 41(2): p. 145–151.

18. Dong, Y., et al., Epidemiology of COVID-19 Among Children in China. Pediatrics, 2020: p. e20200702.

19. Wu, Z. and J.M. McGoogan, Characteristics of and Important Lessons From the Coronavirus Disease 2019 (COVID-19) Outbreak in China: Summary of a Report of 72314 Cases From the Chinese Center for Disease Control, and Prevention. JAMA, 2020.

20. Zhang, J.J., et al., Clinical characteristics of 140 patients infected with SARS-CoV-2 in Wuhan, China. Allergy, 2020.

21. Liu, C., et al., [Preliminary study of the relationship between novel coronavirus pneumonia and liver function damage: a multicenter study]. Zhonghua Gan Zang Bing Za Zhi, 2020. 28(2): p. 148–152.

22. Zhang, J.J., et al., Distinct characteristics of COVID-19 patients with initial rRT-PCR-positive and rRT-PCR-negative results for SARS-CoV-2. Allergy, 2020.

23. Li, X., et al., Risk factors for severity and mortality in adult COVID-19 inpatients in Wuhan. J Allergy Clin Immunol, 2020.

24. Brough, H.A., et al., Managing childhood allergies and immunodeficiencies during respiratory virus epidemics - the 2020 COVID-19 pandemic. Pediatr Allergy Immunol, 2020.

25. Garg, S., et al., Hospitalization Rates and Characteristics of Patients Hospitalized with Laboratory-Confirmed Coronavirus Disease 2019 - COVID-NET, 14 States, March 1–30, 2020. MMWR Morb Mortal Wkly Rep, 2020. 69(15): p. 458–464.

26. Dong, X., et al., Eleven faces of coronavirus disease 2019. Allergy, 2020.

27. Ashraf, M.A., et al., COVID-19 in Iran, a comprehensive investigation from exposure to treatment outcomes. medRxiv, 2020: p. 2020.04.20.20072421.

28. Auld, S., et al., ICU and ventilator mortality among critically ill adults with COVID-19. medRxiv, 2020: p. 2020.04.23.20076737.

29. Barrett, E.S., et al., Prevalence of SARS-CoV-2 infection in previously undiagnosed health care workers at the onset of the U.S. COVID-19 epidemic. medRxiv, 2020: p. 2020.04.20.20072470.

30. Bello-Chavolla, O.Y., et al., Predicting mortality due to SARS-CoV-2: A mechanistic score relating and diabetes to COVID-19 outcomes in Mexico. medRxiv, 2020: p. 2020.04.20.20072223.

31. Burn, E., et al., An international characterisation of patients hospitalised with COVID-19 and a comparison with those previously hospitalised with influenza. medRxiv, 2020: p. 2020.04.22.20074336.

32. Carr, E., et al., Supplementing the National Early Warning Score (NEWS2) for anticipating early deterioration among patients with COVID-19 infection. medRxiv, 2020: p. 2020.04.24.20078006.

33. Clark, A., et al., How many are at increased risk of severe COVID-19 disease? Rapid global, regional and national estimates for 2020. medRxiv, 2020: p. 2020.04.18.20064774.

34. Cummings, M.J., et al., Epidemiology, clinical course, and outcomes of critically ill adults with COVID-19 in New York City: a prospective cohort study. medRxiv, 2020: p. 2020.04.15.20067157.

35. Ni Lochlainn, M., et al., Key predictors of attending hospital with COVID19: An association study from the COVID Symptom Tracker App in 2,618,948 individuals. medRxiv, 2020: p. 2020.04.25.20079251.

36. Paranjpe, I., et al., Clinical Characteristics of Hospitalized Covid-19 Patients in New York City. medRxiv, 2020: p. 2020.04.19.20062117.

37. Qian, Z., et al., Between-centre differences for COVID-19 ICU mortality from early data in England. medRxiv, 2020: p. 2020.04.19.20070722.

38. Vaid, A., et al., Machine Learning to Predict Mortality and Critical Events in COVID-19 Positive New York City Patients. medRxiv, 2020: p. 2020.04.26.20073411.

39. Whitman, J.D., et al., Test performance evaluation of SARS-CoV-2 serological assays. medRxiv, 2020: p. 2020.04.25.20074856.

40. zhang, h., et al., Potential Factors for Prediction of Disease Severity of COVID-19 Patients. medRxiv, 2020: p. 2020.03.20.20039818.

41. Team, C.C.-R., Coronavirus Disease 2019 in Children - United States, February 12-April 2, 2020. MMWR Morb Mortal Wkly Rep, 2020. 69(14): p. 422–426.

42. Shen, K., et al., Diagnosis, treatment, and prevention of 2019 novel coronavirus infection in children: experts’ consensus statement. World J Pediatr, 2020.

43. COVID-19: GINA answers to frequently asked questions on asthma management. 2020 3/25/2020 05/01/2020]; Available from: https://ginasthma.org/covid-19-gina-answers-to-frequently-asked-questions-on-asthma-management/.

44. Iramain, R., et al., Salbutamol and ipratropium by inhaler is superior to nebulizer in children with severe acute asthma exacerbation: Randomized clinical trial. Pediatr Pulmonol, 2019. 54(4): p. 372–377.

45. BTS advice for healthcare professionals treating patients with asthma. 2020 05/01/2020]; Available from: https://www.brit-thoracic.org.uk/document-library/quality-improvement/covid-19/bts-advice-for-healthcare-professionals-treating-patients-with-asthma/.

46. CDC. COVID-19: People who are at higher risk - people with asthma. 2020 04/02/2020 05/01/2020]; Available from: https://www.cdc.gov/coronavirus/2019-ncov/specific-groups/asthma.html.

47. Abrams, E., G. Jong, and C. Yang. Paediatric asthma and COVID-19. 2020 04/01/2020 05/01/2020]; Available from: https://www.cps.ca/en/documents/position/paediatric-asthma-and-covid-19.

48. Shaker, M.S., et al., COVID-19: Pandemic Contingency Planning for the Allergy and Immunology Clinic. J Allergy Clin Immunol Pract, 2020.

49. Wilson, K.C., et al. COVID-19: Interim guidance on management pending empirical evidence. From an American Thoracic Society-led International Task Force. 2020 04/03/2020 05/01/2020]; Available from: https://www.thoracic.org/covid/covid-19-guidance.pdf.

50. Pediatrics, A.A.o. Critical updates on COVID-19. 2020 04/23/2020 05/01/2020]; Available from: https://services.aap.org/en/pages/2019-novel-coronavirus-covid-19-infections/.

51. Halpin, D.M.G., et al., Do chronic respiratory diseases or their treatment affect the risk of SARS-CoV-2 infection? Lancet Respir Med, 2020.

52. Yamaya, M., et al., Inhibitory effects of glycopyrronium, formoterol, and budesonide on coronavirus HCoV-229E replication and cytokine production by primary cultures of human nasal and tracheal epithelial cells. Respir Investig, 2020.

53. Matsuyama, S., et al., The inhaled corticosteroid ciclesonide blocks coronavirus RNA replication by targeting viral NSP15. bioRxiv, 2020: p. 2020.03.11.987016.

54. Jackson, D.J., et al., Association of Respiratory Allergy, Asthma and Expression of the SARS-CoV-2 Receptor, ACE2. J Allergy Clin Immunol, 2020.

55. Prompetchara, E., C. Ketloy, and T. Palaga, Immune responses in COVID-19 and potential vaccines: Lessons learned from SARS and MERS epidemic. Asian Pac J Allergy Immunol, 2020. 38(1): p. 1–9.

